# Analysis of online information available for treatment of depression

**DOI:** 10.1101/19008284

**Authors:** Zachary Clarke, Pietro Ghezzi

## Abstract

The Internet has become a prime source of health information available to the public. Our aims were to assess the content and quality of online information for the treatment of depression.

We searched, “How to cure depression” on Google and analysed the first 200 websites according to the website typology and a standard health information assessment tool, the Journal of the American Medical Association (JAMA) criteria (presence of authorship, date, disclosure, and references). We also analyzed content in terms of treatments mentioned and developed a Quality Indicator Score (QIS) based on the guidelines for treating depression from the UK National Institute for Health and Care Excellence (NICE).

News websites were the most frequent typology followed by health portals, non-profit, professional and government; commercial websites were the least represented. In the top ten websites, news and health portals remained first and second respectively. Antidepressants were the most mentioned treatment, followed by psychotherapy, lifestyle & exercise, social support, diet. The least mentioned interventions were sunlight & light therapy, routine, and ketamine & psychedelics. Commercial websites preferentially mentioned supplements, while ketamine and psychedelic drugs were the most covered by news outlets. Analysing webpages according to our QIS showed the median of NICE recommended treatments was 2.5 out of 5 possible treatments: antidepressants, lifestyle & exercise, psychotherapy, social support and ECT. Government websites had the highest QIS, news and commercial websites the lowest. Webpages with high QIS ranked higher in Google.

## Introduction

Depression is the leading cause of ill-health and disability worldwide and a major contributor to the overall global burden of disease. It affects about three million people in the UK, yet it has been estimated that only about one-third of these will receive treatment [1].Barriers, such as social stigma [2, 3] and delayed access to health care, result in a higher percentage usage of the Internet by patients with depression, as it is an anonymous and instant source of widely available information. For instance, a study revealed as many as 79% of young adults with mental health challenges seek information about their conditions online [4]. With such websites being searched for, often by vulnerable individuals, the content and quality of such online health information gleaned is of great import, in terms of its reliability, accuracy, completeness, safety, bias and validity [5].

This study aims to analyse the content and quality of information available online for treatment of depression which would be found by the public when searching the Internet. To do so we have applied a workflow that we have used previously for other search topics, analyzing the first 200 websites returned by Google® upon searching “how to cure depression”. To evaluate the health information quality (HIQ) of the pages returned, we used the JAMA criteria as a standard assessment tool for their trustworthiness [6]. The JAMA score is based on the presence of four intrinsic criteria of a webpage (authorship, date, references and ownership of the website) and does not take into account the scientific validity of the content. As a proxy indicator of scientific correctness, we looked at the content of each website to identify types of treatments and interventions described. We then assessed whether the information was evidence-based using the guidelines of the UK National Institute of Clinical Excellence (NICE) as a reference [7, 8].

## Methods

### Data collection

We searched the phrase, “How to cure depression” in Google.com from Brighton, UK, using the Google Chrome browser in September 2018. Prior to beginning the search, we cleared all browsing, caches and cookie histories to minimize personalization of the results. The first 200 results returned on the SERP were saved on a spreadsheet. We chose the sample size of 200 as this has previously been suitable for identifying composition differences in the SERP [9, 10].

The workflow of the analysis is reported in Figure 1. Of the 200 websites returned by Google, 26 were excluded. Exclusion criteria were: links not working (n=3), links with no content (n=1), pages requiring subscription or payment (n=3), audio or video (n=11), books (n=7); typically dynamic pages generated by amazon.com), pages about veterinary medicine (n=1). Thus, 174 webpages were included in the analysis.

**Figure 1.**
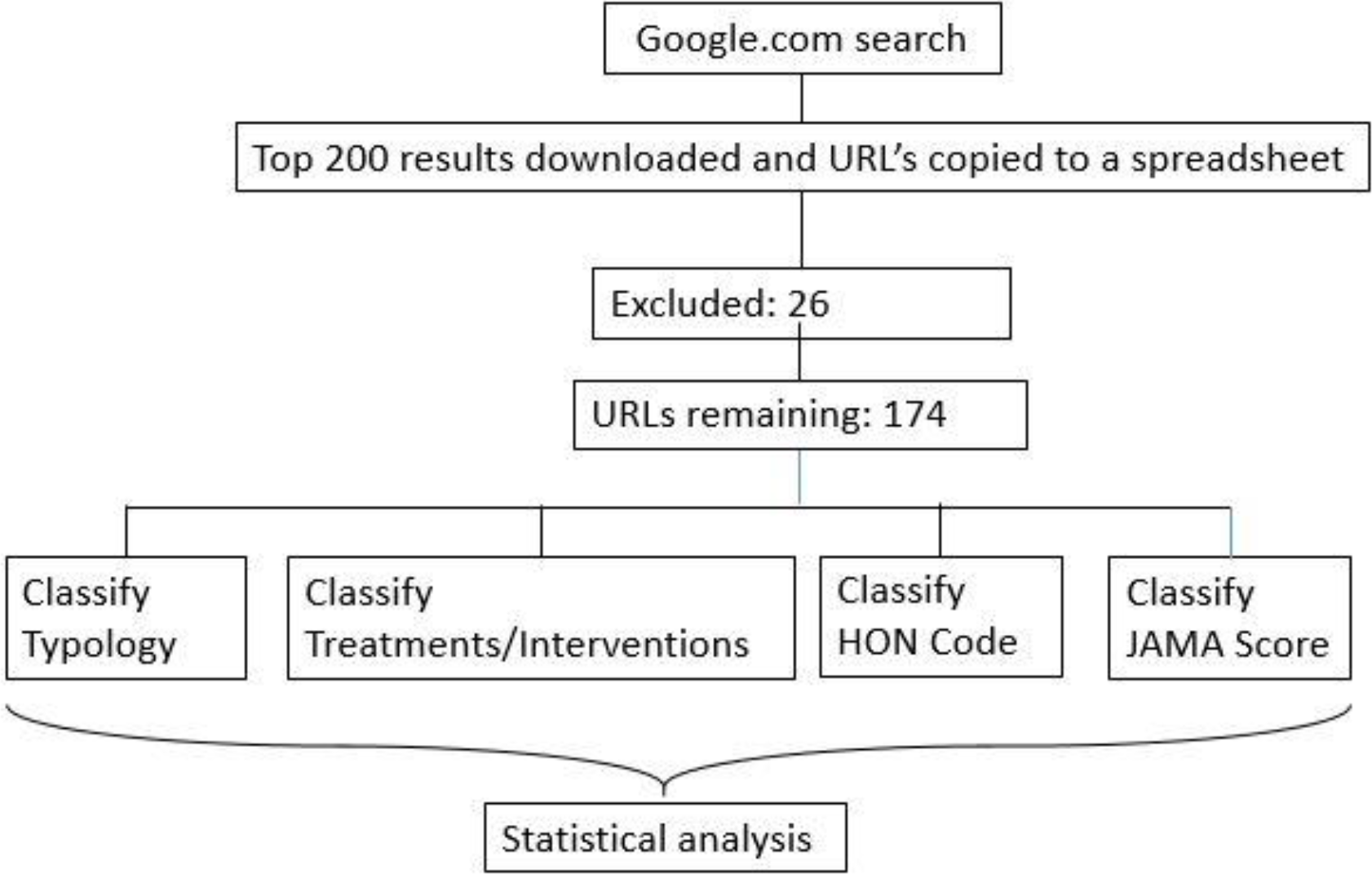
Flowchart of data collection and analysis.

### Website classification

Websites were then visited and classified to one of the following typologies: commercial (C), government (G), health Portal (HP), news (N), non-Profit (NP) or professional (P). Those not fitting into any typology were classified as others (O). Examples of this classification are provided in Supplementary Table 1. The classification was done by two independent researchers and disagreements were resolved with discussion. The agreement was 93% and its strength is considered very good (inter-rater agreement was: Kappa=0.907; confidence interval from 0.857 to 0.958).

### Content analysis

The content was then analyzed and the type of intervention or treatment mentioned was recorded; webpages often mentioned more than one treatment or intervention. The list of intervention was compiled while going along with the analysis.

### Health information quality indicators

The JAMA score was calculated for each webpage as described [6]. Briefly, we noted whether the page contained the following: 1) author; 2) date of writing or update; 3) external references; and 4) owner of the website. Each of these scored “1” if the information was present on the page or within three clicks from the page, according to the three-click rule [11]. Therefore, the JAMA score ranges from 0 to 4.

We also searched for the presence of the Health on the Net (HON) code, a quality seal obtained by the HON foundation and takes into account various criteria of transparency and trustworthiness [12].

### Statistical analysis

When two frequencies were compared, a two-tailed Fisher’s exact test was used. When comparing frequencies in more than two groups, P values were corrected for multiple comparison using the Benjamini-Hochberg procedure to acquire a false discovery rate of 0.25.

The JAMA score and any other quality score in two groups were compared using a two-tailed Mann-Whitney test. Multiple comparisons used a Kruskal-Wallis test followed by Dunn’s multiple comparison test.

When quantifying the agreement in assigning website typology, inter-rater agreement between two raters was quantified as Cohen’s kappa based in Fleiss’ equations [13].

All analyses were performed using GraphPad Prism 7.0 for Windows (GraphPad Software, Sand Diego, CA, USA). Inter-rater agreement was calculated using GraphPad QuickCalc.

## Results

### Type of websites returned

Websites from news outlets were the most frequent, followed by health portals, and the trend was similar for the whole search and in the top 10 websites (Figure 2).

**Figure 2.**
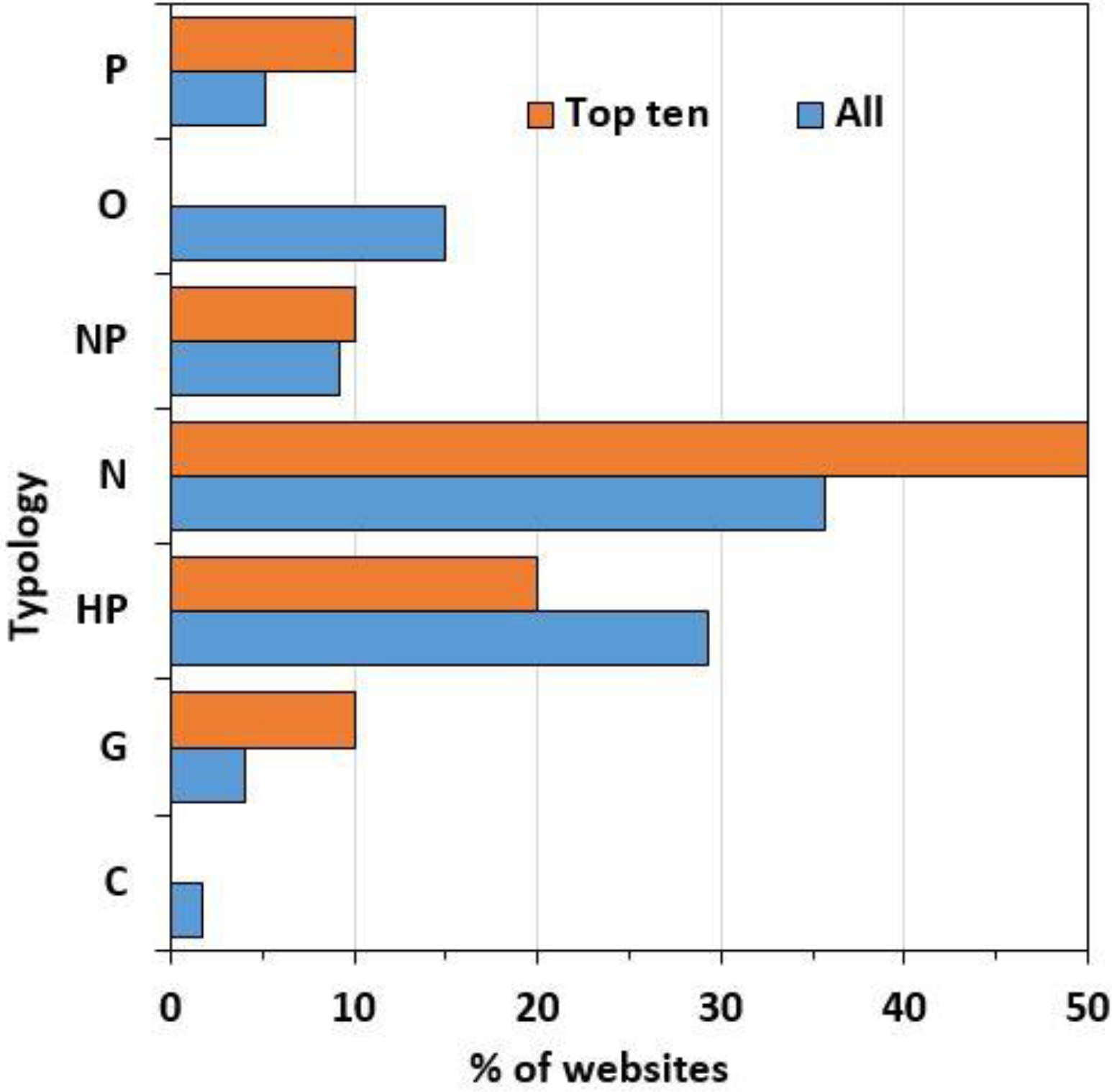
Distribution of websites by typology. Data indicate the percentage of websites overall (blue, n = 174) and in the top ten results (orange, n = 10).

### Health Information Quality Indicators

The median JAMA score of the 174 websites analyzed was 3, IQR: [2, 3]. Figure 3 shows the frequency distribution (A) and analysis of the JAMA score across different typologies (B). Of all websites, 119 (68%) had a JAMA score ≥ 3, which is considered good [13]. The JAMA score varied across typologies with health portals showing the highest values while commercial and government pages had the lowest. The value for health portals was significantly higher when compared with commercial, government, news or no profit. There was no significant difference in the JAMA score of the top ten webpages and those ranked 11-200 by Google (top 10, median 3, IQR [2,4]; all other webpages, median 3, IQR [0,3].

**Figure 3.**
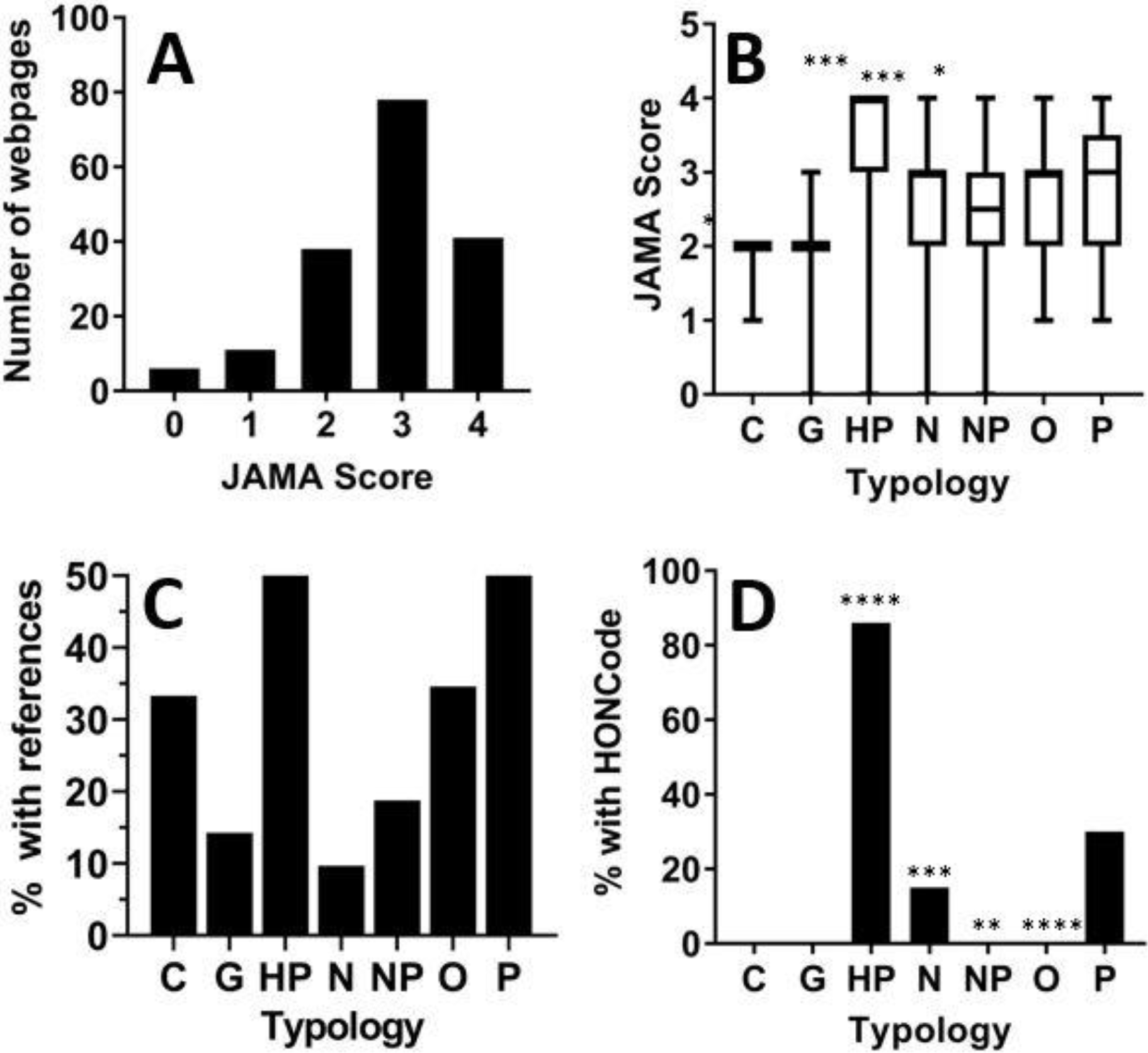
JAMA Score and HONcode in websites of different typologies. Panel A: Frequency distribution of JAMA score in all 174 websites. Panel B: JAMA score of websites of different typology; data are median, IQR, minimum, maximum. *P<0.05, ****P<0.001 vs. HP by a two-tailed Kruskal-Wallis test followed by Dunn’s multiple comparison test. Panel C: Percentage of webpages having external references for the information provided. Panel D: Percentage of webpages in each typology displaying the HONCode seal. **P<0.01, ***P<0.001; ****P<0.0001 vs. all other typologies combined by Fisher’s test (statistical significance was confirmed after correction for multiple comparison using the Benjamini-Hochberg procedure with a false discovery rate set at 5%).

When looking at the percentage of webpages that provided external references as evidence for the information provided, one of the JAMA criteria, the condition was met by 64 websites (37% of total). However, as shown in Figure 3C, there was a huge difference across typologies, ranging from 10% in news webpages to 55% in professional websites and 76% in health portals.

The HONcode seal was displayed by 56 or 174 webpages (32%). As shown in Figure 3D, only health portals, news and professional websites displayed a HONcode. When the percentage in each typology was compared with the remaining websites (not with the total search to avoid comparing overlapping data), health portals had a significantly higher frequency of HONcode certification while news, no profit and other websites had a significantly lower one.

There was no significant difference in the presence of HONcode in the top ten webpages and that in those ranked 11-200 by Google (not shown)

### Content analysis: interventions

Figure 4 shows the interventions described and their frequency in the total search (174 webpages) and in the top ten pages returned by Google. Antidepressants, psychotherapy and lifestyle & exercise were the most mentioned treatments. The least mentioned interventions were sunlight & light therapy, routine, and ketamine & psychedelics. However, in the top ten webpages, psychotherapy and diet were the most frequently mentioned treatment mentioned (Figure 4, orange bars).

**Figure 4.**
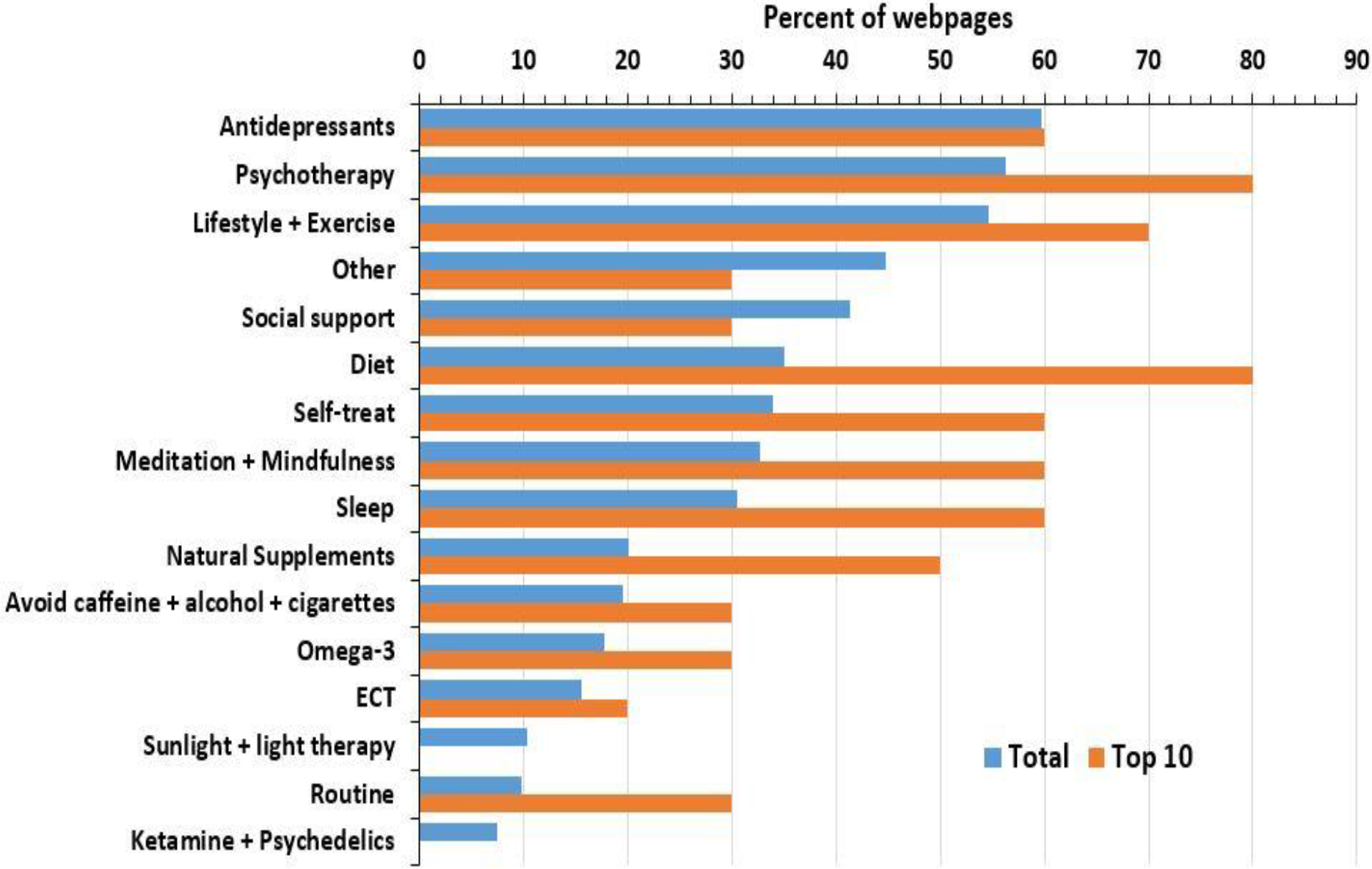
Interventions mentioned. Data show the percentage of interventions mentioned in the top ten (orange, n = 10) and overall (blue, n = 174). As most websites mentioned more than one treatment or intervention, the total percentages add up to more than 100%.

As “completeness” may be considered a criterion of information quality [5, 14] we analyzed the number of interventions described by webpages. The median number of interventions mentioned by webpages was 4, IQR [2,7], with a minimum of 0 and a maximum of 13. Interestingly, webpages with more complete information seems to rank better. The number of interventions mentioned in the top ten webpages (median, 6, min, 2, max, 9, IQR, 5.75-8.25, n=10) was significantly higher compared to the remaining websites (median, 4, min, 0, max, 13, IQR, 2-7, n=164. P<0.05 by a two-tailed Mann-Whitney’s test). Figure 5 shows how many indications were mentioned in each typology of websites. Government and no-profit organizations’ websites had the highest median while health portals and news had the lowest. In a multiple comparison, the value for news was significantly lower compared to government and health portals.

**Figure 5.**
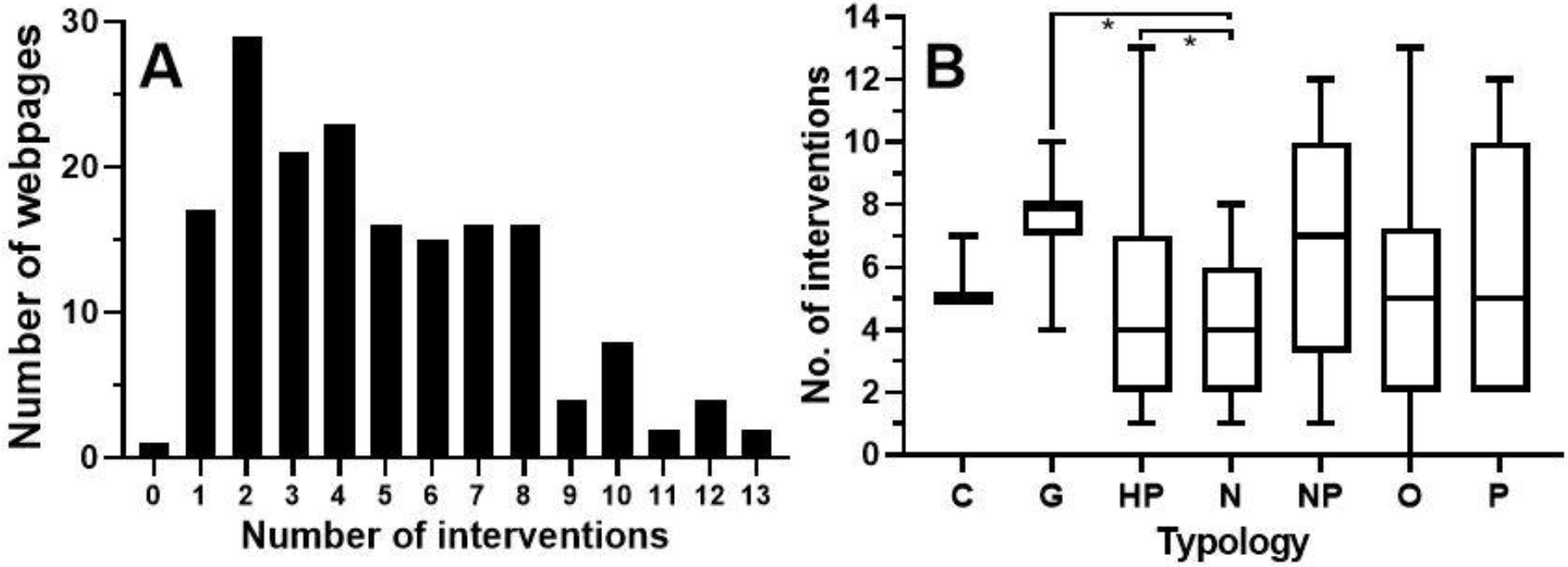
Number of interventions mentioned by websites of different typology. Panel A: Frequency distribution of QIS across all 174 webpages. Panel B: Number of interventions in webpages of different typology; Data are median, IQR, minimum, maximum. *P<0.05 by a two tailed Kruskal-Wallis test followed by Dunn’s multiple comparison test.

Finally, we investigated whether certain website typologies were more inclined to discuss certain treatment types while mentioning others less. The frequency of interventions mentioned across the different website typologies is shown in Figure 7, along with the expected frequency, observed in the total search. Noticeably, commercial websites had a positive bias towards supplements. Ketamine and psychedelic drugs were preferentially mentioned by news websites (of the 13 webpages mentioning it, 10 were from news websites).

**Figure 6.**
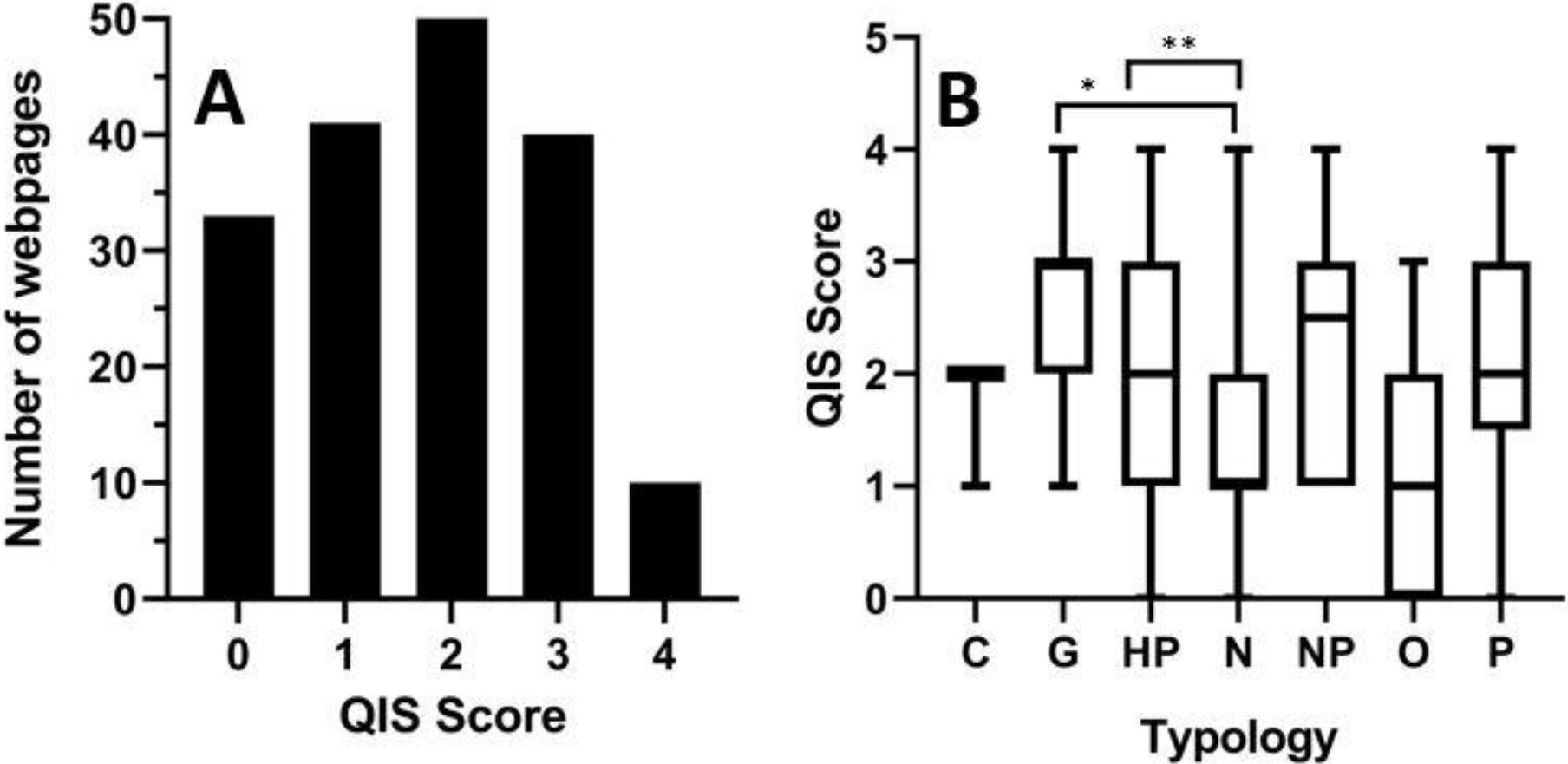
Quality indicator score of webpages. Panel A: Frequency distribution of QIS across all 174 webpages. Panel B: QIS of webpages of different typology; data are median, IQR, minimum, maximum. *P<0.05, **P<0.01 by a two tailed Kruskal-Wallis test followed by Dunn’s multiple comparison test.

**Figure 7.**
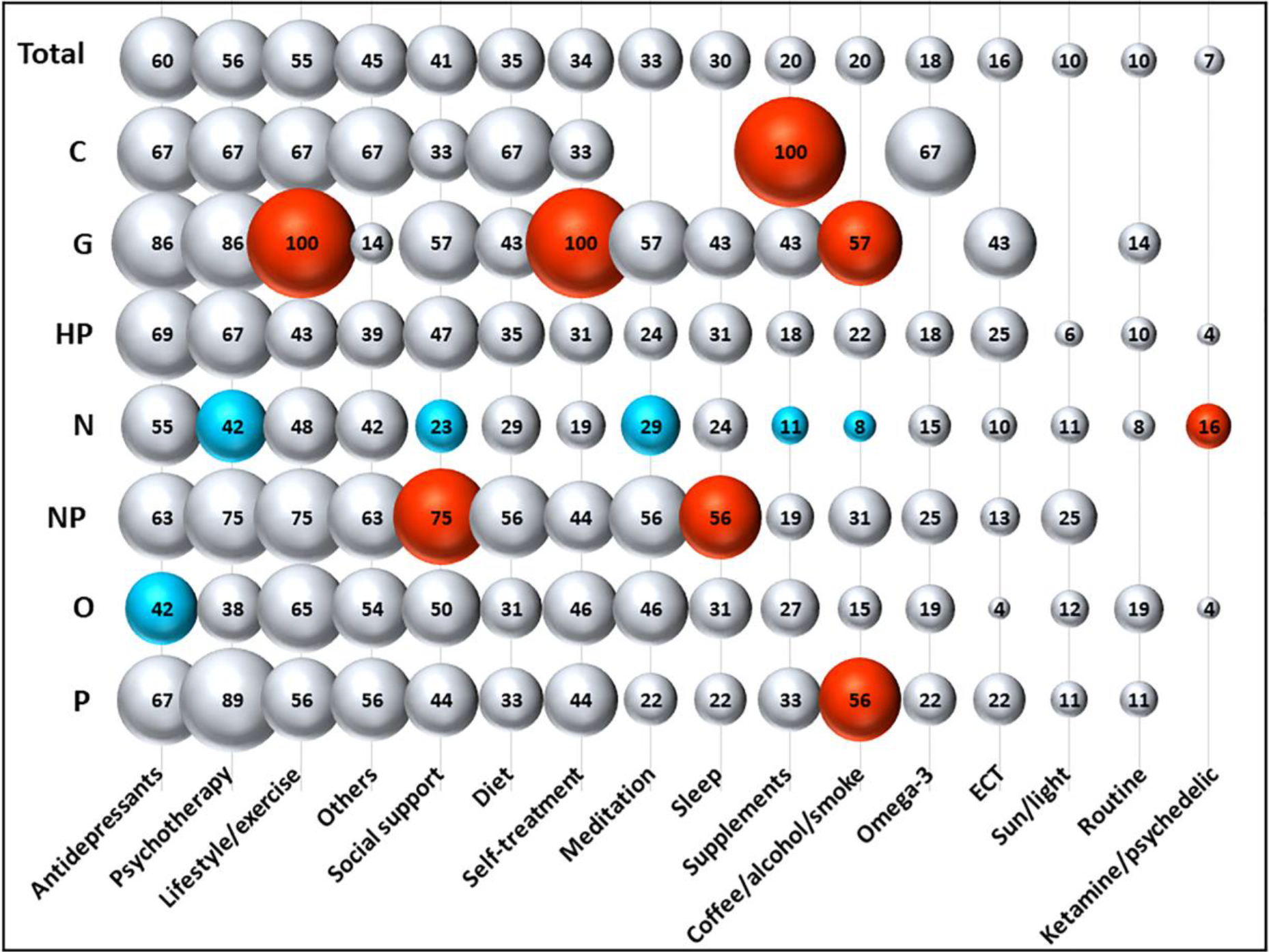
Mention of different types of intervention across typologies of websites. Data are expressed as a percentage of the total number of websites mentioning a type of intervention. Data labels indicate the percentage of websites in a typology that mention a specific intervention. Red and blue indicate an intervention that is significantly over- or under-represented, respectively, in a typology compared with the expected frequency (in the overall SERP). Statistical significance (P<0.05) was calculated comparing the observed frequency and the expected one by Fisher’s test. This was followed by Benjamini-Hochberg correction for multiple comparisons with a false discovery rate of 0.25.

### A quality indicator score (QIS) based on evidence-based medicine

The proposed QIS looks at whether a webpage mentions interventions for depression that are evidence-based as a proxy for “scientific correctness” of the information. We have used the NICE guidelines on treating depression [7, 8]. These include five types of intervention: antidepressants, lifestyle/exercise, psychotherapy, social support and ECT. A webpage was then scored from 0 to 5 depending on how many of these treatments were mentioned. The QIS of all webpages ranged from 0 to 4, with a median of 2, IQR [1,3]. As shown in Figure 6A, over 40% of the webpages mentioned one or less evidence-based intervention. There was no difference between the top 10 webpages and the rest of the SERP (not shown).

As seen in Figure 6B, government websites displayed the highest median QIS, and news websites the lowest. Multiple comparison of all the website typologies against each other showed that the differences between QIS for news vs government and news vs non-profit were statistically significant. Since news websites were the largest represented typology overall and in the top ten websites, we compared the QIS of news (median, 2, min, 0, max, 5, IQR, 1-3, n=62) with that of the remaining webpages (median, 3, min, 0, max, 5, IQR, 1.25-4, n=112) and the difference was statistically significant (P<0.001 by a two-tailed Mann-Whitney’s test).

## Discussion

Our study, which analysed 174 websites, provides a current and comprehensive picture of the quality and content of health information accessible online about treatments for depression. We also performed a sub-analysis of the top ten websites to determine whether there were any significant differences in the visibility of information obtained by users. The observation that news was the largest typology of websites (35% of the whole SERP and 50% of the top ten) indicates the newsworthiness of this topic. Our results confirms a previous study which found a substantial increase in newspapers coverage on mental health between 2008 and 2014 in England [15]. More critically, research has shown news is a primary and influential source of information for sufferers with depression and other mental illnesses [16, 17]. A study conducted by MIND concluded that news coverage has a direct impact on people suffering from mental illness disorders with 50% of sufferers stating that media coverage had a negative impact on their own mental health and 34% feeling more depressed as a result [18].

A possible reason for the high percentage of news websites observed in this study may paradoxically be an indication of the increased drive by the government to remove the stigma associated with depression and promote a better public awareness and understanding of mental health issues [19]. Such campaigns have all impacted on the increased news publicity of depression and other mental health issues [15].

Our HIQ analysis study found that although news had a high median JAMA score of 3 out of 4, the commonly missing JAMA criterion was “referencing”, which was present in just 10% of news, indicating a lack of evidence-based information. This is confirmed by the fact that new websites scored the lowest QIS. We observed, as previous studies have concluded, that content tended to focus on a single news worthy topic [20, 21], rather than providing well-rounded information about depression or its available treatments [22]. Where treatments were mentioned, there was a bias towards over-mentioning of ketamine & psychedelics and under-mentioning of standard treatments like psychotherapy or social support, which are potentially not as newsworthy as the use of ketamine as a “wonder drug”.

Health portals ranked second largest (nearly 30%) of website typologies. In accordance with previous papers [23], our analysis found that health portal websites had the majority of HONcode certifications. Although one limitation with HONcode certification is that accuracy is not measured, studies have found certified educational material is more accurate [24]. Health portals also scored the highest median JAMA score of 4 and QIS score of 3, confirming they had a higher than average quality of information and thus good evidence-based health information coverage.

We have attempted to develop an evidence-based scoring system allowing a more accurate review and analysis of the quality of health information content provided by websites. It is based on a gold standard set of treatments and interventions which we used as a comparison against treatments mentioned by websites. However, one limitation is that ECT, one of the five treatments in the NICE care pathway, is not mentioned very often as it is a specialist extreme case.

Our study found commercial websites made up only three out of 174 websites and was not represented at all in the top ten. This is in line with other studies on breast cancer treatment and influenza prevention where commercial websites were similarly ranked low [23, 25]. This could, however, be due to the fact that companies selling nutritional supplements and various alternative medicine products do not promote these for the treatment of depression. Due to the extremely low number of commercial websites, the results are effectively insignificant and could potentially be ignored as impacting on sufferers.

A limitation of our study is that of search results change with time, search history and location. Additionally, although clearing all cookies and browsing histories prior to our study to avoid search biases, our geographical location was based on using google.com and so the SERP would not have displayed websites in other languages. We performed a sub-analysis of the top ten websites on the assumption that users are unlikely to open more than the first ten websites returned on the SERP [26], but it is also recognised from studies using eye-tracking devices that another limitation is that users may not necessarily open websites in visibility order but instead randomly [27, 28].

Another limitation is that information quality depends not only on the purpose for which information was provided, but also on how it is used [29]. QIS and completeness are valid only for patient-oriented webpages and generic searches - as the search term was “how to cure depression”. Obviously, if the search string was specifically referring to a treatment (such as serotonin reuptake inhibitors) it would be expected to have webpages only mentioning that treatment.

## Data Availability

We attach a supplementary file containing the raw data in a spreadsheet

---

**Supplementary Table 1.** Examples of website typologies.

| **Typology** | **Description** | **Examples** |
| --- | --- | --- |
| Commercial (C) | A website that aims to sell products or services for a profit. | www.depression-helper.com |
| Government (G) | A website of a government body. | www.nhs.uk |
| Health Portal (HP) | A website that provides information on health or health-related issues. | www.webmd.com<br>www.healthline.com |
| News (N) | A website for distribution of news. | www.independent.co.uk |
| Non-Profit (NP) | A website that does not make a profit but whose purpose is to provide charitable, supportive and educational services. | www.helpguide.org<br>www.mind.org.uk |
| Other (O) | A website that does not fall into any other category. | www.quora.com<br>wikipedia.org |
| Professional (P) | A website created by professional health providers. | www.mayoclinic.org |

